# Diagnosis of Schizophrenia and its Subtypes Using MRI and Machine Learning

**DOI:** 10.1101/2024.08.09.24311726

**Authors:** Hosna Tavakoli, Reza Rostami, Reza Shalbaf, Mohammad-Reza Nazem-Zadeh

## Abstract

**Purpose:** The neurobiological heterogeneity present in schizophrenia remains poorly understood. This likely contributes to the limited success of existing treatments and the observed variability in treatment responses. Our objective was to employ magnetic resonance imaging (MRI) and machine learning (ML) algorithms to improve the classification of schizophrenia and its subtypes.

**Method:** We utilized a public dataset provided by the UCLA Consortium for Neuropsychiatric Research, containing structural MRI and resting-state fMRI (rsfMRI) data. We integrated all individuals within the dataset diagnosed with schizophrenia (N=50); along with age- and gender-matched healthy individuals (N=50). We extracted volumetrics of 66 subcortical and thickness of 72 cortical regions. Additionally, we obtained four graph-based measures for 116 intracranial regions from rsfMRI data including degree, betweenness centrality, participation coefficient, and local efficiency. Employing conventional ML methods, we sought to distinguish the patients with schizophrenia from healthy individuals. Furthermore, we applied the methods for discriminating subtypes of schizophrenia. To streamline the feature set, various feature selection techniques were applied. Furthermore, a validation phase involved employing the model on a dataset domestically acquired using the same imaging assessments (N=13). Finally, we explored the correlation between neuroimaging features and behavioral assessments.

**Finding:** The classification accuracy reached as high as 79% in distinguishing schizophrenia patients from healthy in the UCLA dataset. This result was achieved by the k-nearest neighbor algorithm, utilizing 12 brain neuroimaging features, selected by the feature selection method of Minimum Redundancy Maximum Relevance (MRMR). The model demonstrated high effectiveness (85% accuracy) in estimating the disease vs. control label for a new dataset acquired domestically. Using a linear SVM on 62 features obtained from MRMR, patients with schizophrenic subtypes were classified with an accuracy of 64%. The highest spearman correlation coefficient between the neuroimaging features and behavioral assessments was observed between degree of the postcentral gyrus and mean reaction time in the verbal capacity task (r = 0.49, p = 0.001).

**Conclusion:** The findings of this study underscore the utility of MRI and ML algorithms in enhancing the diagnostic process for schizophrenia. Furthermore, these methods hold promise for detecting both brain-related abnormalities and cognitive impairments associated with this disorder.

**Highlights:**

- The neurobiological heterogeneity present in schizophrenia remains poorly understood.
- This likely contributes to the limited success of existing treatments and the observed variability in treatment responses.
- Magnetic resonance imaging (MRI) and machine learning (ML) algorithms can improve the classification of schizophrenia and its subtypes.
- Structural and functional measures of MRI can discriminate Schizophrenia form healthy individuals with almost 80% accuracy.
- Paranoid is the most distinguishable subtype of schizophrenia.

## 1 Introduction

Schizophrenia is a serious mental health disorder that affects feelings, thoughts, and behavior. There are complications and heterogeneities, which have made its treatment less effective. The diagnosis for schizophrenia mostly relies on self-reports, behavioral observations, and psychiatric history, which have led to an average response to the antipsychotic medications as the mainstream treatment (de Araujo et al., 2012). In a systematic review of 101 studies, the treatment-resistant patients exhibit malfunction in the dopaminergic system and hypersensitivity to dopamine level in comparison with patients responding to antipsychotic treatment (Iasevoli et al., 2023).

Magnetic Resonance Imaging (MRI) as a neuroimaging tool has been a great help to explore the neural basis of psychiatric disorders including schizophrenia. Introducing new biomarkers based on MRI findings is so promising that it is suggested as an add-on diagnosis method for schizophrenia (Galderisi et al., 2019). Another promising field in which MRI has been helpful is personalized medicine. With the pieces of evidence MRI brought to the field, adjusting the parameters of treatments such as brain stimulation based on individual features draws some attention (Zangen et al., 2023, Klooster et al., 2022, Cole et al., 2022). Capturing differences in structure of brain between healthy and schizophrenic patients using MRI (Zhao et al., 2022, Li et al., 2022, Brenner et al., 2022) as well as the function (Zhu et al., 2022, Saris et al., 2022, Scognamiglio and Houenou, 2014, Zeng et al., 2022), is prompted scientists to invest more on this modality. The MRI modalities are capable to discriminate healthy from schizophrenia patients, for instance a simple linear model on voxel-based morphometry features can diagnose sufficiently, even on data from different sites and several scanners (Nemoto et al., 2020). A review also highlights that neuroimaging studies in schizophrenia revealed the significant role of drug abuse in the loss of brain volume of patients (Walter et al., 2012). Employment of brain function and structure simultaneously as well as their interaction can strongly examine schizophrenia patients from healthy individuals (Antonucci et al., 2022).

MRI studies on brain structures revealed that the ventricular volume is associated with poor treatment outcome in patients with schizophrenia (Lieberman et al., 2001). Moreover, studying brain morphology in schizophrenia has proven that the treatment-resistance patients are in more progressive stages of changes in brain morphology than treatment-responsive cases (Sone et al., 2023). Decreased thickness of cortical regions such as the insula and superior temporal gyrus has been also reported in first-episode drug-naïve schizophrenics compared to healthy controls (Song et al., 2015). In a diffusion tensor imaging (DTI) study, schizophrenia patients with severe hallucination showed disintegrated fiber integrity in the connection between frontal and temporoparietal language area (de Weijer et al., 2011). In another DTI study, white matter abnormalities in frontal, parietal and temporal regions were found associated with a poor treatment outcome (Mitelman and Buchsbaum, 2007, Molina et al., 2008). Enlargement of white matter volumes was also observed in treatment-resistance patients compared to treatment-responsive patients (Molina et al., 2008, Anderson et al., 2015).

Despite many efforts, there are investigation in the field to find prognostic biomarkers and identify treatment-resistance cases with schizophrenia in order to offer a proper treatment at early stages (Jiao et al., 2022, Vita et al., 2019). With the significant advancement of technology, there is more optimism for introducing innovative and objective methodologies, which may aid in a better understanding of the heterogeneity of schizophrenia and suggestion of a potent individualized treatment.

Functional connectivity in brain as an identification of spontaneous interaction of regions obtained during resting-state obtained abnormalities in favor of schizophrenia. By exploring the resting-state fMRI (rsfMRI) of schizophrenic patients with auditory hallucinations, a hypoconnectivity between the primary auditory cortex and secondary auditory cortical regions was found (Gavrilescu et al., 2010). Various measures extracted from rsfMRI can project different aspects of schizophrenia effects on the brain. For example, abnormal functional connectivity in schizophrenia was shown in individual regional homogeneity (ReHo), the amplitude of low-frequency fluctuations (ALFF), and the degree centrality values extracted from rsfMRI (Li et al., 2023). There are benefits in applying graph analyses on functional connectivity in order to characterize the brain networks (Rubinov and Sporns, 2010). There is also evidence for the ability of graph measures to capture significant lower segregation and higher integration in structural connectome (Gao et al., 2023, Wang et al., 2017).

Moreover, some studies point to MRI’s ability to distinguish between subgroups of patients with schizophrenia which can explain a portion of heterogeneities in this disorder. Structural MRI has been used to distinct between schizophrenic subtypes, namely a morphometry study suggesting a reduction in cortical folding in disorganized subtypes of schizophrenia relative to healthy controls, predominantly manifested in the left hemisphere of the paranoid subtype (Sallet et al., 2003). Patients over the course of schizophrenia revealed significant aberration in cortical thickness (Zhao et al., 2022). In a multisite study, subgrouping schizophrenia using clustering approaches on brain structures has resulted in three distinct groups with different cognitive functions (Xiao et al., 2022). A valuable study supporting neurobiological differences between paranoid and non-paranoid schizophrenia (Lutz et al., 2020), identified larger bilateral hippocampi, right amygdala, and their subfield volumes in paranoids compared to non-paranoid cases. It supports that structural MRI can play a major role diagnosis of schizophrenic subtypes.

The combination of MRI with machine learning (ML) offers a new tool to exploit novel biomarkers, diagnose illnesses, and forecast the response to a particular treatment in a more accurate manner as a result of the development of new mathematical algorithms and data collecting technologies. To find patterns and traits connected to schizophrenia, ML algorithms can be trained to examine huge volumes of MRI data from numerous patients. This will facilitate the development of tailored treatment programs and more precise diagnostic decision-making by clinicians. By applying several ML models, the researchers identified some pre-treatment clinical measures to predict the treatment outcome in depression (Webb et al., 2020). The outcome of antipsychotic medications is variable across the patients with schizophrenia. ML algorithms have been shown capable to predict the treatment outcome for the first-episode drug-naïve schizophrenia patients from the functional connection in superior temporal cortex with an accuracy of 82.5% (Cao et al., 2020). Furthermore, resting-state EEG has shown potential in classifying responders vs. non-responders to the brain stimulation treatment (Ebrahimzadeh et al., 2024).

Modalities neuroimaging with ML models works has elevated the accuracy of diagnosis for mental health disorders (Quaak et al., 2021, Wang et al., 2017). However, the number of studies with utilizing ML for subtyping the patients is limited.

The primary objective of this study is to apply ML and MRI to classify patients with schizophrenia and its subtypes. We also seek to reach more accurate discrimination of patients from healthy controls as well as schizophrenia subtypes by utilizing the structural and functional features of the brain. To reach the goals, we first extracted structural features and graph measures from T1-weighted image and rsfMRI respectively. Then, using the conventional ML models, we classified patients to schizophrenia and healthy. Different combinations of features were tested on all models to obtain the best model with the best combination of features. We evaluated the performance of the best model in classification of schizophrenia subgroups from healthy controls. As an extra validation, we acquired a new domestic dataset from the patients diagnosed with schizophrenia to assess the selected models on an unseen test data. We used the same procedure on subtypes label to test whether the conventional models and MRI measures are capable of differentiating between subtypes of schizophrenia. For the final step, the correlation of the extracted features with behavior assessments was inspected to uncover some of associations between the brain and behaviors in the patients with schizophrenia.

## 2 Material and method

### 2.1 Main dataset

We used the dataset from UCLA Consortium for Neuropsychiatric Phenomics (https://openneuro.org/datasets/ds000030/versions/1.0.0) consisting neuroimaging and neuropsychological data from healthy individuals and patients with schizophrenia (Poldrack et al., 2016). Neuroimaging data were acquired at the Ahmanson-Lovelace Brain Mapping Center (Siemens version syngo MR B15) and the Staglin Center for Cognitive Neuroscience (Siemens version syngo MR B17) at the University of California, Los Angeles, USA. The parameters for the high-resolution scan were: 4mm slices, TR/TE=5000/34 ms, 4 averages, Matrix=128 × 128. The parameters for MPRAGE were the following: TR=1.9 s, TE=2.26 ms, FOV =250 mm, Matrix =256 × 256, sagittal plane, slice thickness=1 mm, 176 slices. The resting fMRI scan lasted 304 s. Participants were asked to remain relaxed and keep their eyes open; they were not presented any stimuli or asked to respond during the scan.

First, we gathered the information of all 50 schizophrenia patients and then matched them to 50 out of 130 healthy controls by the age and gender The age- and gender matched groups are shown in **Table 1**. We extracted demographics, structural MRI, and resting-state fMRI (rsfMRI) data of both groups. We also used behavioral assessments to investigate their relationships with imaging data. The list of three domains of behavioral tests performed on the subjects is presented in **Table 2** (Poldrack et al., 2016).

**Table 1.** Demographic of healthy and patient groups.

| Group | Number | Age (mean ± std) | Male/Female |
| --- | --- | --- | --- |
| Healthy | 50 | 36.40 ± 8.87 | 38/12 |
| Schizophrenia | 50 | 36.46 ± 8.87 | 38/12 |
| Negative | 26 | 36.69 ± 8.38 | 19/7 |
| Positive | 23 | 36 ± 9.71 | 18/5 |
| Paranoid | 21 | 38.38±8.54 | 17/4 |
| Undifferentiated | 10 | 34.4±8.85 | 8/2 |
| Residual | 6 | 33.66±8.29 | 6/0 |
| Schizoaffective | 11 | 37±8.57 | 4/7 |
| Local dataset (Healthy) | 20 | 31.43±8.34 | 10/10 |
| Local dataset (Patients) | 13 | 33.84±11.58 | 11/2 |

**Table 2.** Behavioral assessments in three domains: Traits, neurocognitive, and neuropsychological.

| Domains | Measures |
| --- | --- |
| **Traits** | Barratt Impulsiveness Scale (BIS-11)<br>Dickman Functional and Dysfunctional Impulsivity Scale<br>Multidimensional Personality Questionnaire (MPQ)—Control subscale<br>Scale for Traits that Increase Risk for Bipolar II Disorder<br>Golden & Meehl's Seven MMPI Items Selected by Taxonomic Method<br>Hypomanic Personality Scale (HPS)<br>Chapman Scales (Perceptual Aberrations, Social Anhedonia, Physical Anhedonia)<br>Temperament and Character Inventory (TCI)<br>Munich Chronotype Questionnaire (MCTQ) |
| **Neurocognitive Tasks** | Task-switching Task (TS)<br>Spatial Capacity Task (SCAP)<br>Verbal Capacity Task (VCAP)<br>Delay Discounting Task (DDT)<br>Balloon Analog Risk Task (BART)<br>Attention Network Task (ANT)<br>Continuous Performance Go/NoGo Task (CPT)<br>Stroop Color Word Task (SCWT)<br>Stop Signal Task (SST)<br>Scene Recognition Task<br>Remember-Know Task (RK)<br>Spatial Maintenance and Manipulation Task (SMNM)<br>Verbal Maintenance and Manipulation Task (VMNM) |
| **Neuropsychological Assessment** | California Verbal Learning Test (CVLT-II)<br>WMS-IV Symbol Span<br>WMS-IV Visual Reproduction<br>WAIS-IV Letter Number Sequencing<br>WMS-IV Digit Span<br>WAIS-IV Vocabulary<br>WAIS-IV Matrix Reasoning<br>Color Trails Test |

We also utilized the Scale for the Assessment of Negative Symptoms (SANS) and Scale for the Assessment of Positive Symptoms (SAPS) to divide patients into Negative and Positive groups. The individuals with negative scores greater than positive ones were put in the Negative; and the ones with positive scores more than negative comprised the Positive group. There were two subjects with equal scores of positive and negative symptoms which were eventually put in the Positive group for the sake of maintaining the balance between the two groups. A further grouping was made based on patients’ subtypes defined by the Structured Clinical Interview for DSM-5 (SCID-5).

### 2.2 Extra validation dataset

For extra validation of ML models to explore how these models would perform on an unseen dataset, 13 patients with schizophrenia along with 20 healthy subjects were recruited with the same imaging and behavioral measurements as the UCLA dataset. The patients were diagnosed by DSM-5 and an MRI session conducted on a 3T MRI system with a 64-channel head coil (Prisma, Siemens, Erlangen, Germany) at the National Brain Mapping Laboratory located at Tehran University, Iran, while attending a neurologist (N. T.) throughout the scans. Each session included a T1-weighted image with following protocol: TR=1.9 s, TE=2.26 ms, FOV = 250 mm, Matrix =256 × 256, Sagittal plane, Slice thickness=1 mm, Resolution= 1 x 1 x 1 mm, 176 slices. The resting-state scan lasted 396 s using the following parameters: TR=1.2 s, TE=30 ms, FOV=192 mm, Matrix = 64 × 64, Sagittal plane, Slice thickness=3 mm, Resolution= 3 x 3 x 3 mm, 42 slices.

### 2.3 Data Harmonization

To reduce the impact of using different scanners, we harmonized the data using ComBat method (Johnson et al., 2006). Empirical Bayesian was used as the Bayesian inference in this method using which, the distribution of latent variables was inferred. We applied the ComBat for both main and extra validation datasets.

### 2.4 Feature extraction

Details of the acquisition parameter and assessments of the UCLA dataset are available in the data descriptor (Poldrack et al., 2016). The data was preprocessed by FMRIPREP version 0.4.4 (http://fmriprep.readthedocs.io). Cortical thickness and subcortical volume were calculated by FreeSurfer v6.0.0 (http://surfer.nmr.mgh.harvard.edu). The structural measures were extracted after motion correction, intensity correction, Talairach registration, normalization, skull stripping, and segmentation. The cortical surface and subcortical volumes were segmented and labeled into 68 and 45 regions (34 for each hemisphere), respectively (Gorgolewski et al., 2017).

The preprocessing of the rsfMRI was performed using a toolbox for Data Processing and Analysis of Brain Imaging (DPABI), which evolved from the Data Processing Assistant for Resting-State fMRI (DPARSF) (Yan et al., 2016). We removed the first 10 slices and then slice timing correction, realignment, brain extraction, and co-registration of the functional image on T1 were done as preprocessing. Then the time series of 116 regions of the AAL atlas (Tzourio-Mazoyer et al., 2002) was calculated for both healthy and patient subjects, for each a matrix with a dimension of 320×116 was generated. We then calculated a 116×116 functional connectivity matrix (an undirected brain network) using Pearson’s correlation coefficient between each pair of time series, and extracted these values as imaging features.

Among the vast measures of brain networks, the centrality graph measures including the degree, betweenness centrality, and participation coefficient were extracted to Local efficiency was also calculated to measure the segregation and the presence of densely interconnected brain networks.

These measures were calculated as follows (Rubinov and Sporns, 2010):

- **Degree** is the number of links connected to a node. Degree of a node *i* is defined as:

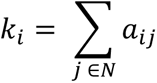

where *N* is the set of all nodes in the network and *a*_*ij*_ is the connection status between nodes *i* and *j*.

- **Betweenness centrality** of node *i* is:

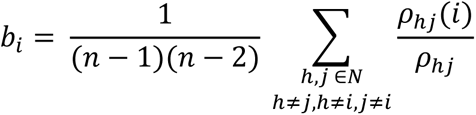

where *ρ*_ℎ*j*_ is the number of shortest paths between ℎ and *j*, and *ρ*_ℎ*j*_(*i*) is the number of shortest paths between ℎ and *j* that pass through *i*.

- **Participation coefficient** of node *i* is:

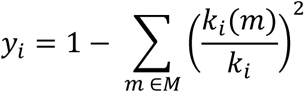

where *M* is the set of modules, and *k*_*i*_(*m*) is the number of links between *i* and all nodes in module *m*. Modularity of a network is *Q* = ∑_*u*∈*M*_[*e*_*uu*_ − (∑_*v*∈*M*_ *e*_*uv*_)^2^], where the network is fully subdivided into a set of nonoverlapping modules *M*, and *e*_*uv*_ is the proportion of all links that connect nodes in module *u* with nodes in module *v*.

- **Local efficiency** of the network is defined as:

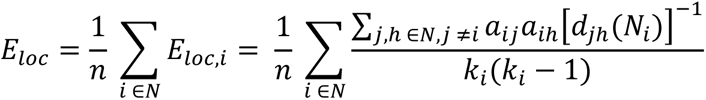

where *E*_*loc*,*i*_ is the local efficiency of node *i*, and *d*_*j*ℎ_(*N*_*i*_) is the length of the shortest path between *j* and ℎ, which contains only neighbors of *i*. We extracted these features for both the datasets in this study.

### 2.5 Statistical analyses

We ran 2-sample t-test to explore differences in MRI measures between the healthy controls and schizophrenia subjects. The Bonferroni correction was used to address the multiple hypothesis testing issues. Since the independent variables (MRI measures) outnumbered the observations, the repeated measure analyses of variance (ANOVA) were conducted to indicate whether or not there are any significant differences between healthy controls, Negative, and Positive groups in the extracted features. We considered the brain region as a repeated factor. There was also another repeated measure ANOVA test to answer the same question about the subtypes of schizophrenia and healthy subjects. The subtypes are Disorganized, Paranoid, Undifferentiated, Residual, and Schizoaffective.

### 2.6 Classification and feature selection

All the procedures of classification and feature selection were operated in MATLAB ver. 2020b. The Conventional ML model including Support vector machine (SVM) with polynomial and linear kernel, k-nearest neighborhood (kNN), Linear Discriminant Analyses (LDA), Linear Regression (LR), Random Forest (RF), and Naïve Bayes (NB) were applied to classify the defined groups. The 10-fold cross-validation approach was executed with 10-time repeats and the performance of the model was measured by calculating the average of mean accuracy of folds among the repeats. Furthermore, three feature selection methods were implemented to reduce the feature dimensions as well as to improve the model’s accuracy. The feature selection methods were:

- **Sequential Forward Selection (SFS)** in which features are sequentially added to an empty candidate set until the addition of further features does not decrease the criterion.
- **Minimum Redundancy Maximum Relevance (MRMR)** is an approach to select features with a high correlation with output (class) and a low correlation with other features in the dataset.
- **Neighborhood Component Analysis (NCA)** is a method for selecting features with the goal of maximizing the prediction accuracy of regression and classification algorithms. It learns the feature weights using a diagonal adaptation of NCA with a regularization term.

SFS is sensitive to the feature sequence so that different arrangement of features results in different sets of final selected features. To address this issue, we implemented SFS 5 times and each time we shuffled the MRI measures before using SFS. The reported accuracy for SFS is the average of 5 repeats.

For MRMR and NCA these steps were performed: 1) apply the method on the feature set, 2) train the ML model with the best feature, 3) add features one by one and replicate the training, 4) determine the features with the highest accuracy. This process was run to find the best model with the lowest feature dimension by using MRMR and NCA.

We performed these steps for classifying the schizophrenia from healthy and also on schizophrenia subtypes. For evaluation, we applied the best model on Negative and Positive groups as well as new unseen dataset including new healthy and patient subjects.

### 2.7 Behavioral and imaging correlation

We inspected the relationship between the imaging features and behavioral assessments. First, we obtained differences between healthy subjects and schizophrenic patients in both imaging and behavioral data. Then, we investigated whether there is any association of MRI measures with behavioral scales. To reduce the sensitivity to the outliers, we utilized the Spearman coefficient method.

## 3 Results

### 3.1 Data drop-out

We dropped out a patient from the Negative and Positive grouping due to the missing SANS and SAPS scores. Furthermore, we dropped the Disorganized subtype for insufficient sample size (N=1).

### 3.2 Statistical results

There were no significant differences in age between any comparative groups based on either the t-test (for two groups) or one-way ANOVA (for more than two groups) (**Table 1**). A two-sample t-test with Bonferroni correction suggested no significant difference in any of structural and graph measures between healthy controls and schizophrenic patients.

There was a significant interaction between group and MRI measures after the Greenhouse–Geisser corrected ANOVA in healthy, Negative, and Positive groups, (*F*(601,1202) = 2.96, *p* = 0.025). Post-hoc analyses using multiple comparison tests revealed significant differences between healthy and Positive groups (*p* = 0.011). Two-sample t-test also identified the significant features as the volume of the right hemisphere, left hemisphere, and the whole cortex.

Another repeated measure ANOVA on healthy controls and subtypes of schizophrenia obtains a significant effect of group on MRI measures after the Greenhouse–Geisser correction (*F*(601,2404) = 2.51, *p* = 0.015). The post hoc results suggested that the differences between residual and healthy groups were the most significant (*p* = 0.007).

### 3.3 Classification results

#### 3.3.1 Healthy and Schizophrenia groups

MRI preprocessing and feature extraction provided a vector with 602 features for each subject including sixty-six subcortical volumes (of 45 subcortical regions plus 21 whole-brain, white matter, and right and left hemisphere cortex), 72 cortical measurements (68 left and right regions plus 4 whole-brain cortical thickness) and 4 graph measures of 116 brain regions (602 = 66 + 72 + 4×116). The accuracy of models is shown in **Figure 1**. The combination of all three sets of imaging measures suggested the best accuracy of 67% using RF classifier. As it is observed, there is an improvement after applying feature selection methods, with the best accuracy 0f 79% achieved by kNN when applied on the 12 featured selected by MRMR. The most important features obtained from MRMR were: thickness of middle temporal and middle frontal gyrus in left hemisphere and insula in right hemisphere, degree of right superior frontal gyrus, the volumes of right hippocampus, right postcentral gyrus, and midline of vermis, participation coefficient of left cuneus and right palladium, betweenness centrality of left postcentral gyrus and left superior frontal gyrus and local efficiency of middle frontal gyrus. Confusion matrix, sensitivity, and specificity of the kNN model with selected features to evaluate the model are in **Table 3**. A high sensitivity reported for schizophrenia group means that the classifier has the ability to designate the individual with disease as positive. The specificity is showing an acceptable false positive result for healthy and schizophrenia groups. The details on other performances and accuracies are available in Table S1 in supplementary.

**Figure 1.**
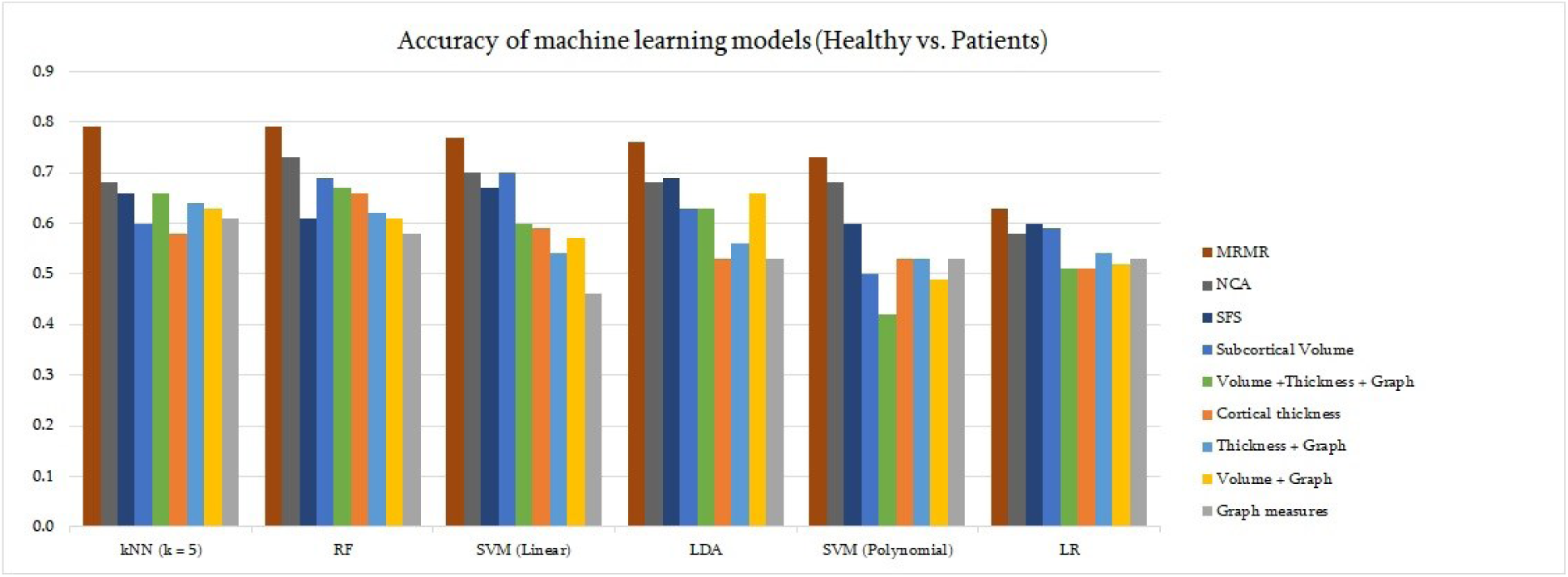
Performance of machine learning models for differentiating schizophrenia vs. healthy with different sets of features. There are six models with nine sets of features. The highest accuracy (79%) belongs to kNN and MRMR, considered as the best model. Although the combination of RF and MRMR resulted in the same accuracy as the combination of kNN and MRMR (79%), the latter combination was chosen because of a lower number of features (12 < 22).

**Table 3.** Confusion matrix of the best model on classifying heathy from schizophrenia with the sensitivity and specificity of the model.

|  |  | True value |  | Sensitivity | Specificity |
| --- | --- | --- | --- | --- | --- |
|  |  | Healthy | Schizophrenia |  |  |
| Prediction | Healthy | 42 | 14 | 0.84 | 0.71 |
|  | Schizophrenia | 8 | 35 | 0.81 | 0.75 |

#### 3.3.2 Performance of selected model on other groups

The validation of the selected model on other group classifications is assessed in this section. **Table 4** are showing the accuracy of the kNN model in classifying each group. The 12 features used in this classification are the same as those extracted from MRMR mentioned in the previous section. The worth performance on the UCLA dataset belongs to healthy, Negative, and Positive groups classification with 51% accuracy. On the other hand, the kNN model with 12 features was able to discriminate healthy subjects from the Positive group with an acceptable accuracy of 74%. After harmonization of the extra validation dataset, although predicting labels of the patients seems a great success with an accuracy of 72%, the standard deviation is high (35%). Prediction of new healthy and patient subjects after harmonization was not noteworthy (58%).

**Table 4.** Accuracy (mean±std) of kNN for classifying the different groups from UCLA dataset and local dataset (13 patients) using 12 features extracted from MRMR method.

| Groups | Accuracy<br>(kNN on 12 features from MRMR) |
| --- | --- |
| Healthy, Negative, Positive | 0.51±0.02 |
| Healthy, Negative | 0.72±0.02 |
| Healthy, Positive | 0.74±0.02 |
| Negative, Positive | 0.52±0.04 |
| Healthy, Schizophrenia (local dataset) | 0.58±0.04 |

#### 3.3.3 Schizophrenia subtypes

There was a drastic inequality between the numbers of samples for each subtype, as shown in **Table 1Error! Reference source not found.**. Four subtypes: Paranoid, Undifferentiated, Residual, and Schizoaffective were considered. The same procedure was adopted as the classification of patients vs. healthy controls. **Figure 2** shows the performance of 7 ML models and 9 sets of features on classifying the subtypes. The highest accuracy of 64% derived from SVM with linear kernel on 62 features obtained from MRMR. The performance of subtype classifier was found inferior compared to the patient vs. control classifier (See Table S2 of supplementary for more details). **Table 5** compares the selected model performance for each group in a confusion matrix form with sensitivity and specificity values. By identifying 14 out of 21, this classifier was the most accurate in differentiating Paranoid subtype with an accuracy of 67%, followed by the Schizoaffective subtype with an accuracy of 64%. The highest sensitivity and specificity in diagnosis of Schizoaffective confirms the great differences of this subgroup with schizophrenia subtypes, with a support towards the most distinguished subtype which is Paranoid.

**Figure 2.**
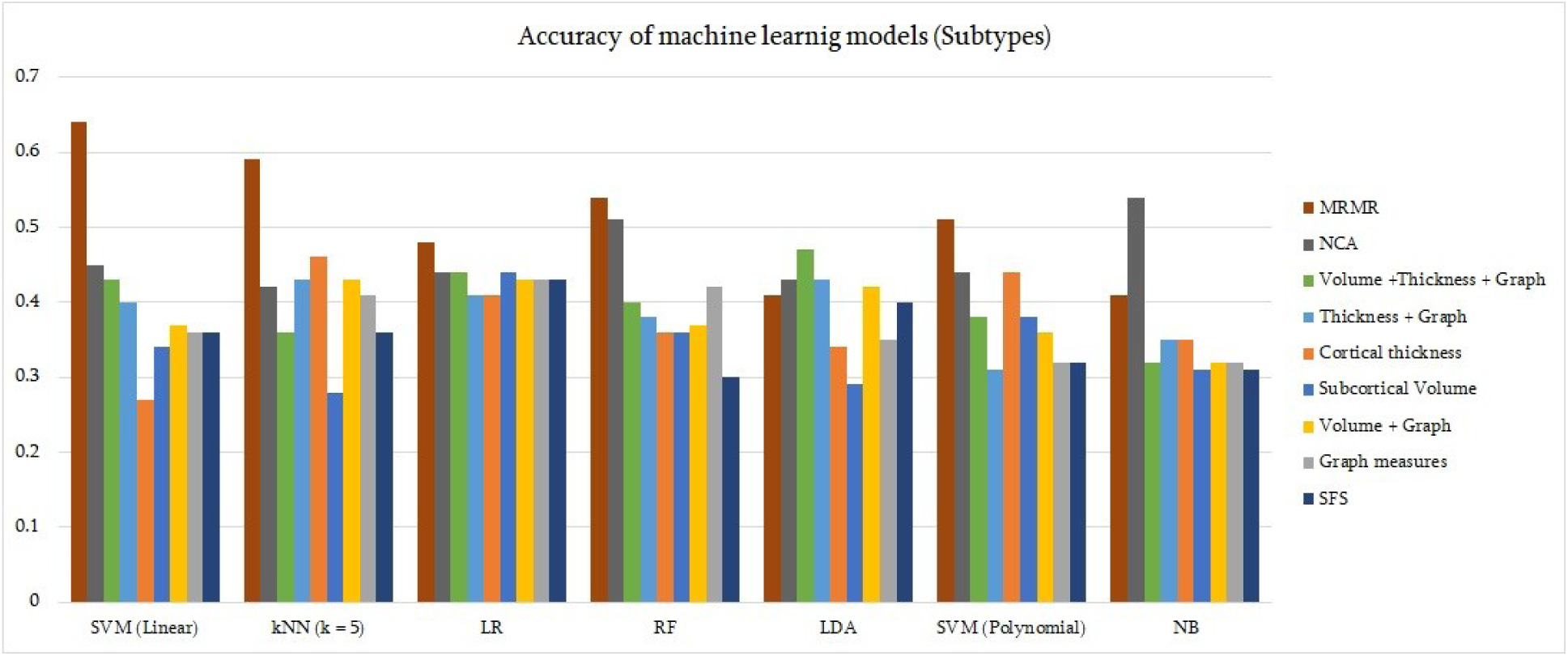
Accuracy of machine learning models and sets of features for differentiating schizophrenic subtypes. SVM with linear kernel on 62 features extracted using MRMR method reached the highest accuracy (64%) of classification.

**Table 5.** Confusion matrix of the best model on classifying the subtypes of schizophrenia with the sensitivity and specificity of the model.

|  |  | True value |  |  |  | Sensitivity | Specificity |
| --- | --- | --- | --- | --- | --- | --- | --- |
|  |  | Paranoid | Undifferentiated | Residual | Schizoaffective |  |  |
| Prediction | Paranoid | 14 | 3 | 2 | 1 | 0.67 | 0.77 |
|  | Undifferentiated | 4 | 6 | 1 | 1 | 0.60 | 0.84 |
|  | Residual | 1 | 1 | 3 | 0 | 0.50 | 0.95 |
|  | Schizoaffective | 2 | 0 | 0 | 8 | 0.80 | 0.95 |

### 3.4 Behavioral Results

**Table 6** lists the behavioral measures with the strongest correlations to each of the 12 imaging features with a significant difference (p < 0.05). The degree of right postcentral and the verbal capacity task showed the highest correlation (r = 0.49, p = 0.001). The thickness of left middle temporal and mean accuracy of manipulation trials in VMNM task showed the second highest positive connection (r = 0.45, p = 0.002). Both the participation coefficient of the left cuneus and the degree of vermis were negatively correlated with the reaction times of two cognitive tasks (r = -0.44, -0.47, p = 0.003, 0.002). The remaining negative correlations (r = -0.42, -0.46, p = 0.005, 0.002) were seen between two MRI measures and the recollection process of two tasks. **Figure 3** shows the most positive and negative correlated imaging features and behavioral scales.

**Figure 3.**
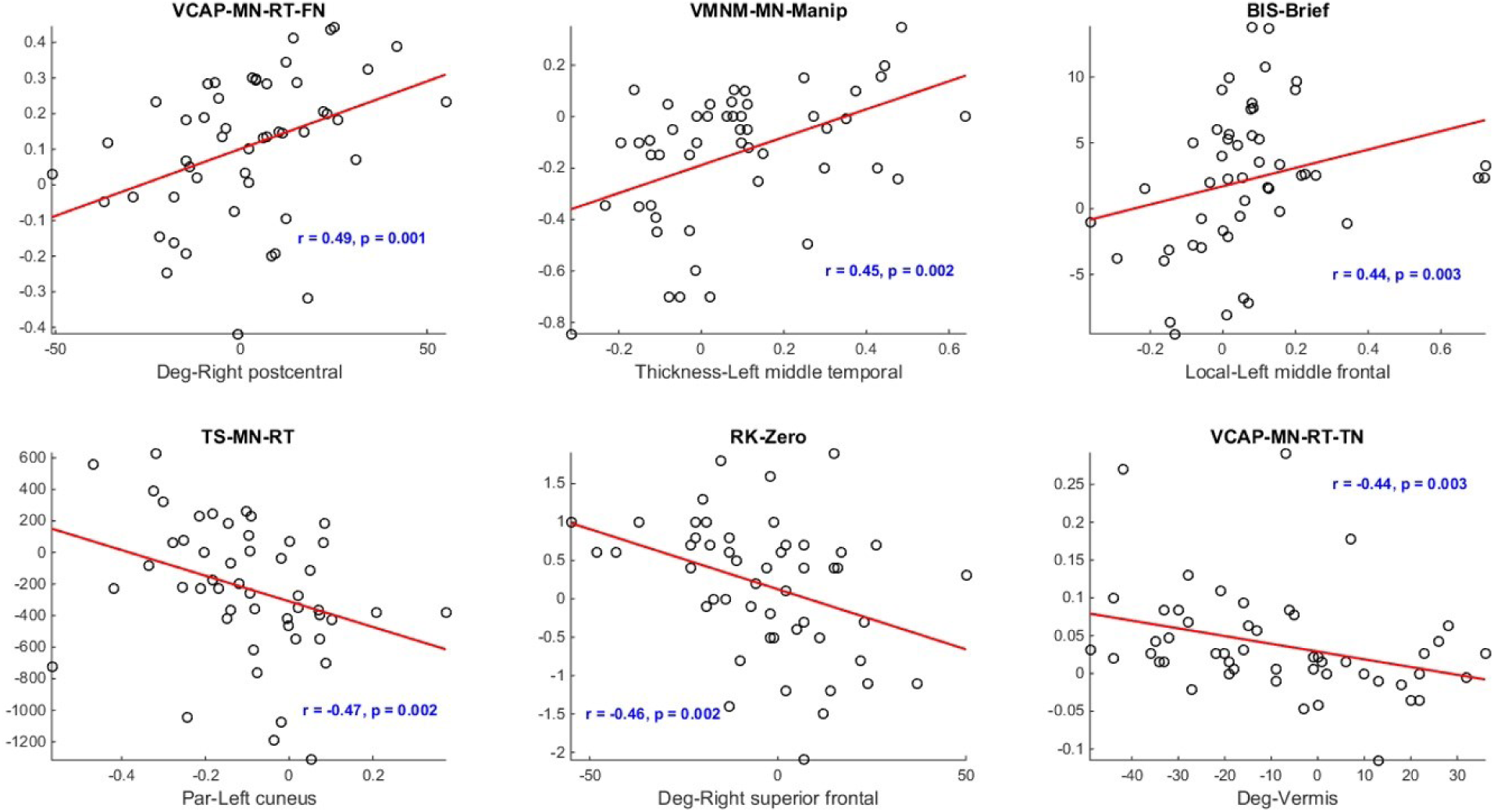
Interaction of the differences observed in 6 extracted MRI measures between the schizophrenia and healthy cohorts, in conjunction with the most closely associated behavioral indicators. The red lines in the scatterplots represents the optimal linear regression correlating MRI and behavioral metrics. Spearman correlation results are denoted as ’r’ and the corresponding p-values are presented above each scatterplot, offering insight into the strength and significance of the observed relationships.

**Table 6.** Spearman’s correlation coefficients and p-values between MRI and behavioral measures.

| **Imaging measures** | **Behavioral items** | **r-value** | **p-value** |
| --- | --- | --- | --- |
| **Thickness-Left middle temporal** | VMNM - Mean Accuracy of Manipulation trials | 0.45 | 0.002 |
| **Degree-Right hippocampus** | SCAP - Number of correct answers | 0.41 | 0.005 |
| **Local efficiency-Left middle frontal** | BIS - Brief | 0.44 | 0.003 |
| **Participation coefficient -Right pallidum** | TCI - Novelty | -0.33 | 0.024 |
| **Degree-Vermis** | VCAP - Mean reaction time of true negatives | -0.44 | 0.003 |
| **Participation coefficient-Left cuneus** | TS - Mean reaction time | -0.47 | 0.002 |
| **Degree-Right postcentral** | VCAP - Mean reaction time of false negatives | 0.49 | 0.001 |
| **Betweenness centrality-Left precentral** | RK - Number of Know responses | 0.38 | 0.010 |
| **Thickness-Left middle frontal** | SST - Stop signal reaction time | 0.39 | 0.008 |
| **Thickness-Right insula** | CVLT - Number of correct recall answers | -0.42 | 0.005 |
| **Betweenness centrality-Left superior frontal** | CVLT - Long delay cued recall | 0.41 | 0.006 |
| **Degree-Right superior frontal** | RK - Zero recalls | -0.46 | 0.002 |

## 4 Discussion

Schizophrenia diagnosis is not merely reliant on a singular method; rather, a combination of physical and psychological assessments aids clinicians to achieve accurate diagnoses and treatments. MRI serves as a diagnostic tool, revealing structural and functional brain abnormalities that may distinguish the patients with schizophrenia from healthy individuals. Moreover, recent strides in ML exhibit potential in leveraging MRI data to identify and forecast outcomes in schizophrenia (Rozycki et al., 2017, Yassin et al., 2020). This study offers substantial evidence of ML’s significance in diagnosing and understanding schizophrenia through both structural and functional imaging data. Achieving an accuracy of approximately 80%, the utilized MRI measures including cortical thickness and graph metrics, effectively differentiate between healthy individuals and the patients. This performance on the specific dataset stands as one of the notable accomplishments to date (Quaak et al., 2021, Matsubara et al., 2019). In order to diagnose patients, the suggested strategy by this study needs to extract only 12 features from MRI images. This may be advantageous in reducing the computation cost and model’s complexity. Clinical subtypes of schizophrenia are less noted in the context of classification. According to the results of this study, Paranoid subtype can be discriminated from normal with a decent accuracy (67%). This may be a valuable point to obtain the neural differences of schizophrenia subtypes.

The most pertinent features chosen as significant for classification were graph measures derived from rsfMRI data. The application of graph theory has offered novel insights into the functional connections and the collaborative behaviors of brain regions in the context of human cognitive functions and behaviors (Farahani et al., 2019). Degree, local efficiency, betweenness centrality, and participation coefficient represent graph measures computed from rsfMRI data can provide insights into various facets of brain functional connectivity. Research has demonstrated that the organization of brain networks in individuals with schizophrenia, as identified through graph theoretical analysis, deviates from the typical patterns found in healthy controls (Gao et al., 2023).

Five of the twelve selected features are associated with the attention network including the thickness of the middle frontal gyrus in the left hemisphere and the insula in the right hemisphere, the degree of the right superior frontal gyrus, the betweenness centrality of the left superior frontal gyrus, and the local efficiency of the middle frontal gyrus. These findings are in agreement with existing literature. Conclusions drawn from both imaging data and behavioral observations suggest that attentional deficits in patients manifest in performance on attention-related tasks and are reflected in the brain’s activity and connectivity within the attention network (Jimenez et al., 2016, Roiser et al., 2013, Ioakeimidis et al., 2020).

The presence of nine functional-related measures highlights that distinctions in brain function between patients and healthy individuals were more evident. Conversely, subcortical volume values played a negligible role in discerning patients from healthy subjects. Given that the disorder tends to preserve brain structure, particularly in its early stages, this outcome was foreseeable. Consequently, it can be inferred that MRI measures associated with brain networks might hold the potential to enhance the accuracy of diagnostic procedures.

Another noteworthy finding of this study was that the chosen model exhibited superior performance in distinguishing Positive group from healthy individuals compared to Negative group. This suggests that individuals with positive symptoms show greater deviations from the normal state in terms of brain function and structure, compared to patients with negative symptoms. Substantiating this interpretation, statistical analyses confirmed that alterations in the Positive group significantly impact the overall cortical thickness of the brain. This aligns with prior research indicating distinct neural underpinnings for negative and positive symptoms (Vanes et al., 2019).

In an additional validation step, the selected model displayed robust performance when applied to a new dataset, achieving an accuracy rate of nearly 85%. However, the presence of a high standard deviation suggests that the model’s reliability on previously unseen data might be somewhat compromised.

A multi-class classification task in general faces more challenges than a two-class classification, which is the case for the conventional classification methods used in this study. There is more concentration on clustering strategies as opposed to the classification, for subtyping the schizophrenia using behavioral (Lefort-Besnard et al., 2018, Chen et al., 2020) or anatomical data (Chand et al., 2020). We managed to solve this problem in a certain way through adjustments, supported by the validated performance on the local data acquired in this study. From the results, we observed that all models performed poorly on classifying the subtypes. In addition, the small number of data and the unbalanced distribution of patients in subtype groups has intensified the classification difficulty. This seems to be the reason why studies on subtyping patients are quite limited. However, these issues can be dealt with to a certain degree by using data augmentation approach along with developed ML models.

A significant contribution of neuroimaging data and ML approaches lies within the capacity to unveil associations between brain characteristics and behavioral patterns in psychiatric disorders (Drysdale et al., 2017). Although there are only a few studies investigating that, the results are promising. Schizophrenia patients with low and high social anhedonia were classified based on temporal and spatial networks extracted from fMRI task (Krohne et al., 2019). Deep learning methods on task -based fMRI features suggested the inferior and middle temporal lobe to be sufficiently informative to classify schizophrenia versus healthy subjects (Oh et al., 2019). Another successful deep learning application in diagnosis of schizophrenia has used the structural MRI features and a 3D convolutional neural network architecture (Zhang et al., 2022). The most distinguished regions between control and patients were subcortical cortex and ventricles, pivotal regions in cognitive, affective and social functions. Our results support that the most robust connection pertains to the degree of the postcentral gyrus and the Variable Central Attentional Performance (VCAP) task (r=0.49). A positive association indicates that as the degree of aberration from the norm in the postcentral node increases, the difference in reaction time during the VCAP task between patients and healthy individuals becomes more pronounced. The postcentral gyrus, situated in the parietal lobe, serves as the primary sensory receptive area in the human brain.

Existing literature suggests that working memory deficits are prevalent in numerous psychiatric disorders. A meta-analysis has revealed that the dysfunction of working memory in individuals with schizophrenia may stem from activation abnormalities in regions within the parietal lobe and supplementary motor area—areas closely linked to, though not identical to, the postcentral gyrus (Wu and Jiang, 2020).

We observed a robust negative correlation between the participation coefficient of the right cuneus and reaction time during the switch task underscores the cuneus’s integral role in executive function, and its impairment is discernible in individuals with schizophrenia (Huang et al., 2022, Nyatega et al., 2021). Moreover, the association between the superior frontal gyrus and recall performance in the remember-know task can also be highlighted (Huang et al., 2022).

CVLT test measures episodic verbal learning and memory, particularly in the recall segment. The findings also indicate that performance on the CVLT test declines in schizophrenia patients as insula thickness decreases. There is ample evidence to support the insula’s participation in episodic memory (Vatansever et al., 2021, Dahlgren et al., 2020), and as we have demonstrated, structural alteration in the insula is among the factors that may contribute to cognitive dysfunction in schizophrenia.

Furthermore, we found that alterations in brain structure, specifically cortical thickness, attributed to the mental condition may cause changes in behavior (Ehrlich et al., 2011, Zhao et al., 2022, Fan et al., 2023). Notably, a majority of the behavioral measures linked to imaging metrics fall within the neurocognitive domain, with the exception of the Barratt Impulsiveness Scale (BIS) and the Temperament and Character Inventory (TCI), which belong to the traits domain.

One notable limitation of this study was the relatively small size of the training dataset utilized for the ML model. It’s important to note that employing larger datasets can yield more robust model performance. At present, the availability of a comparable dataset with a substantial volume of MRI data, encompassing both structural and functional aspects of the brain, alongside comprehensive behavioral and cognitive assessments of psychiatric patients—particularly individuals with schizophrenia—is limited. Addressing this challenge might necessitate a collaborative effort across multiple research centers to generate a dataset of sufficient size and diversity, thereby providing more reliable insights to the field.

In cases where such comprehensive datasets do become available, the application of deep learning techniques and neural networks could be explored to more effectively harness the features and achieve enhanced understanding, yielding more refined outcomes.

### Clinical implications

It is a proven state that MRI and fMRI can distinguish the differences in structure and function of the brain between schizophrenia patients and normal individuals. The neuroimaging features along with clinical and behavioral characteristics can determine subtypes of schizophrenia. On the other hand, MRI measures and clinical and behavioral data can be used as features of the input data to be fed into an ML algorithm to learn the subtypes from all types of features together. Then the subtype of a new neuroimaging and behavioral data recorded from an individual can be predicted by this trained ML model which can be negative, positive, or one of the cognitive subtypes (**Figure 4**). This application obtained from our study can pave the way to a new individualized medicine and help the therapeutic approach that targets either positive or negative symptoms, such as add-on TMS or other medicine to regulate symptoms linked to distinct subtypes. The findings of this work can also help in understanding the underlying neural basis of the negative and positive symptoms. To estimate the subtypes, this model could be employed in place of neuropsychological tests with subjective and other patient-related variability. (Carruthers et al., 2019, Gurvich et al., 2023, Dean et al., 2022).

**Figure 4.**
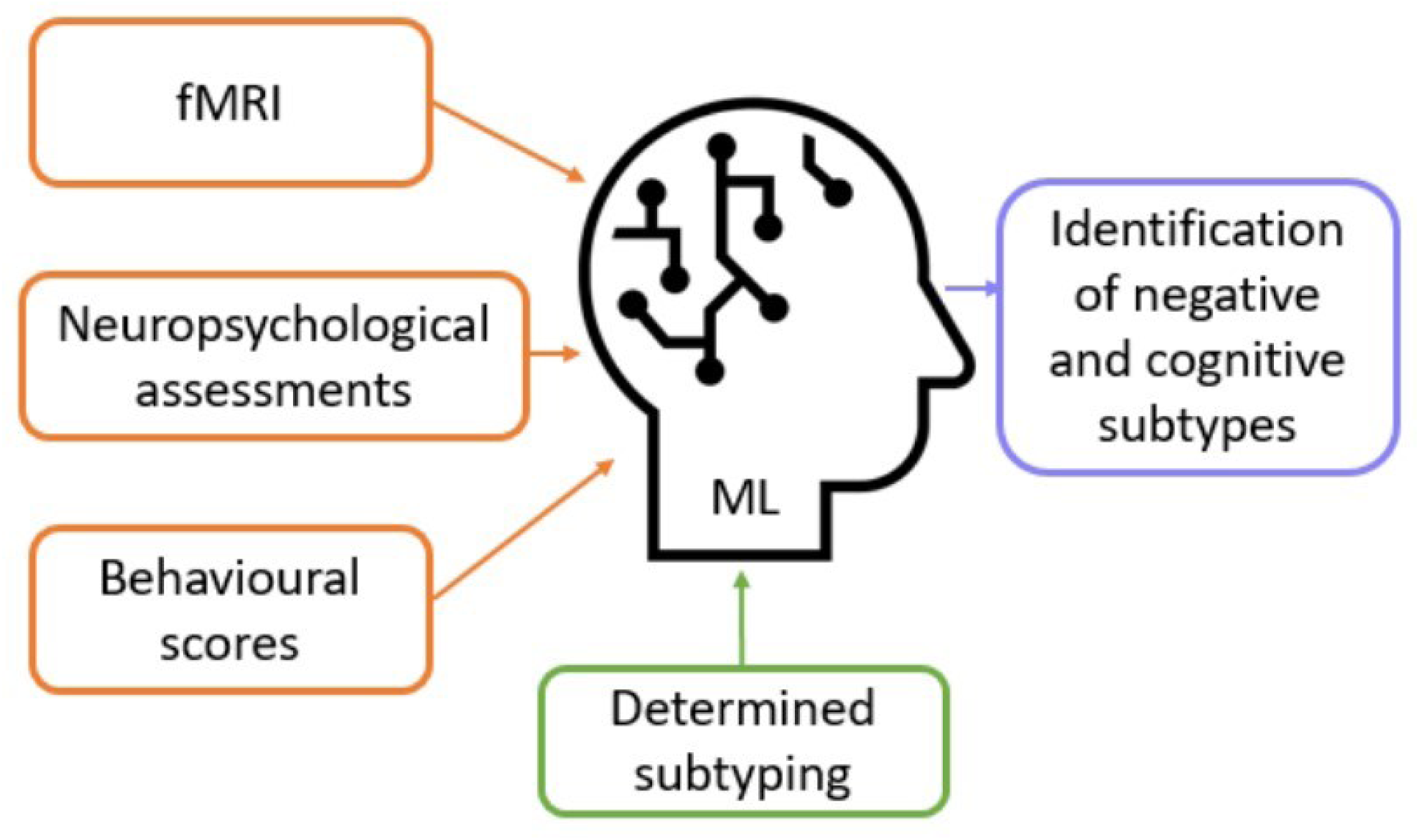
A suggestion for ML model to learn the subtypes of individuals from different types of features.

## 5 Conclusion

This study has effectively classified individuals with schizophrenia and healthy subjects with a commendable level of accuracy, leveraging the structural and functional attributes of MRI data alongside conventional machine learning models. The utilization of graph theory has emerged as a powerful approach in the analysis of functional brain data, offering a comprehensive depiction of various aspects of brain connectivity. Notably, the feature selection process predominantly prioritized graph measures extracted from rsfMRI data, signifying their relevance in the context of this study.

Furthermore, the identification of meaningful correlations between brain characteristics and behavioral manifestations related to schizophrenia aligns harmoniously with existing literature. These outcomes reinforce the notion that the fusion of machine learning methodologies with feature selection techniques holds the potential to unearth novel biomarkers, consequently contributing to the enhancement of diagnosis and treatment strategies for psychiatric disorders.

## Data Availability

All data produced in the present study are available upon reasonable request to the authors

## Supplementary information

The supplementary material can be found in a separate file titled “Supplementary_Material”.

## Acknowledgements

Authors must recognize and show graduate towards the prodigious contribution of Iranian National Brain Mapping Laboratory (NBML), Tehran, Iran, for data acquisition service.

## Declaration

### Ethics approval and consent to participate

All participants provided informed consent according to the study protocol approved by the ethics committee of research, Iran University of Medical Sciences.

### Consent for publication

Not applicable.

### Availability of data and materials

All data can be available by making a proper request to the corresponding author.

### Competing interests

The authors have no competing interests that might be perceived to influence the results and/or discussion reported in this paper.

### Funding

This work was supported by Strategic Technologies Development of National Elite Foundation.

### Authors’ contributions

The authors confirm contribution to the paper as follows: study conception and design, analysis and interpretation of results, draft manuscript preparation: Hosna Tavakoli, Mohammad-Reza Nazem-Zadeh, data collection: Hosna Tavakoli, Mohammad-Reza Nazem-Zadeh, Reza Rostami, review and editing: Mohammad-Reza Nazem-Zadeh, Reza Shalbaf, supervision: Mohammad-Reza Nazem-Zadeh. All authors reviewed the results and approved the final version of the manuscript.

## Notes

### Competing Interest Statement

The authors have declared no competing interest.

