## Supplementray_Material for "Diagnosis of Schizophrenia and its Subtypes Using MRI and Machine Learning"

**Table S1** Accuracy (mean±std) of machine learning models applying different features and combining with feature selection methods for classification of healthy and schizophrenia groups.

| Model / Features | kNN (k = 5) | SVM (Linear) | SVM (Polynomial) | LDA | LR | RF | NB |
|---|---|---|---|---|---|---|---|
| Subcortical Volume | 0.60±0.03 | 0.70±0.02 | 0.50±0.00 | 0.63±0.02 | 0.59±0.02 | 0.69±0.03 | 0.64±0.00 |
| Cortical thickness | 0.58±0.02 | 0.59±0.03 | 0.53±0.01 | 0.53±0.03 | 0.51±0.03 | 0.66±0.03 | 0.69±0.01 |
| Graph measures | 0.61±0.02 | 0.46±0.03 | 0.53±0.01 | 0.53±0.02 | 0.53±0.04 | 0.58±0.02 | 0.61±0.00 |
| Volume + Graph | 0.63±0.04 | 0.57±0.02 | 0.49±0.00 | 0.66±0.01 | 0.52±0.21 | 0.61±0.03 | 0.65±0.01 |
| Thickness + Graph | 0.64±0.02 | 0.54±0.03 | 0.53±0.01 | 0.56±0.02 | 0.54±0.26 | 0.62±0.02 | 0.57±0.02 |
| Volume +Thickness + Graph | 0.66±0.02 | 0.60±0.02 | 0.42±0.01 | 0.63±0.01 | 0.51±0.02 | 0.67±0.03 | 0.64±0.02 |
| SFS (5 features) | 0.66±0.02 | 0.67±0.02 | 0.60±0.02 | 0.69±0.01 | 0.60±0.17 | 0.61±0.04 | 0.66±0.00 |
| MRMR (Number of features) | 0.79±0.04 [1] (12) | 0.77±0.03 (11) | 0.73±0.03 (19) | 0.76±0.07 (6) | 0.63±0.02 (60) | 0.79±0.03 (22) | 0.75±0.02 (23) |
| NCA (Number of features) | 0.68±0.04 (38) | 0.70±0.04 (34) | 0.68±0.03 (39) | 0.68±0.48 (9) | 0.58±0.01 (9) | 0.73±0.05 (44) | 0.64±0.02 (11) |

[1] Highest accuracy

**Table S2** Accuracy (mean±std) of machine learning models applying different features and combining with feature selection methods for classification of healthy and subtypes of schizophrenia.

| Model / Features | kNN (k = 5) | SVM (Linear) | SVM (polynomial) | LDA | LR | RF | NB |
|---|---|---|---|---|---|---|---|
| Subcortical Volume | 0.28±0.03 | 0.34±0.04 | 0.38±0.03 | 0.29±0.03 | 0.44±0.00 | 0.36±0.04 | 0.35±0.02 |
| Cortical thickness | 0.46±0.02 | 0.27±0.04 | 0.44±0.01 | 0.34±0.02 | 0.41±0.01 | 0.36±0.03 | 0.31±0.01 |
| Graph measures | 0.41±0.03 | 0.36±0.02 | 0.32±0.03 | 0.35±0.01 | 0.43±0.00 | 0.42±0.01 | 0.35±0.02 |
| Volume + Graph | 0.43±0.03 | 0.37±0.02 | 0.36±0.04 | 0.42±0.01 | 0.43±0.01 | 0.37±0.03 | 0.32±0.02 |
| Thickness + Graph | 0.43±0.03 | 0.40±0.03 | 0.31±0.04 | 0.43±0.02 | 0.41±0.01 | 0.38±0.05 | 0.32±0.02 |
| Volume +Thickness + Graph | 0.36±0.01 | 0.43±0.03 | 0.38±0.03 | 0.47±0.01 | 0.44±0.00 | 0.40±0.03 | 0.35±0.02 |
| SFS (6 features) | 0.36±0.03 | 0.36±0.03 | 0.32±0.03 | 0.40±0.02 | 0.43±0.00 | 0.30±0.03 | 0.32±0.03 |
| MRMR (Number of features) | 0.59±0.04 (32) | 0.64±0.02 [1] (62) | 0.51±0.04 (70) | 0.41±0.04 (1) | 0.48±0.02 (34) | 0.54±0.04 (55) | 0.31±0.02 (1) |
| NCA (Number of features) | 0.42±0.03 (7) | 0.45±0.04 (21) | 0.44±0.05 (97) | 0.43±0.04 (1) | 0.44±0.01 (10) | 0.51±0.02 (1) | 0.41±0.05 (1) |

[1] Highest accuracy
